# Blood omega-3 fatty acids and death from COVID-19: A Pilot Study

**DOI:** 10.1101/2021.01.06.21249354

**Authors:** Arash Asher, Nathan L. Tintle, Michael Myers, Laura Lockshon, Heribert Bacareza, William S. Harris

**Affiliations:** Samuel Oschin Comprehensive Cancer Institute at Cedars-Sinai Medical Center, Los Angeles, CA; Fatty Acid Research Institute, Sioux Falls, SD; Department of Mathematics and Statistics, Dordt University, Sioux Center, IA; Los Alamitos Medical Center, Los Alamitos, CA; Department of Medical Affairs, Cedars-Sinai Medical Center, Los Angeles, CA; Department of Internal Medicine, Sanford School of Medicine, University of South Dakota, Sioux Falls, SD

## Abstract

Very-long chain omega-3 fatty acids (EPA and DHA) have anti-inflammatory properties that may help reduce morbidity and mortality from COVID-19 infection. We conducted a pilot study in 100 patients to test the hypothesis that RBC EPA+DHA levels (the Omega-3 Index, O3I) would be inversely associated with risk for death by analyzing the O3I in banked blood samples drawn at hospital admission. To have adequate power (>80%) in this pilot study, we pre-specified a significance level of 0.10. Fourteen patients died, one of 25 in quartile 4 (Q4) (O3I ≥5.7%) and 13 of 75 in Q1-3. After adjusting for age and sex, the odds ratio for death in patients with an O3I in Q4 vs Q1-3 was 0.25, p=0.07. Thus, we have suggestive evidence that the risk for death from COVID-19 was lower in those with the highest O3I levels. These preliminary findings need to be confirmed in larger studies.

## 1. INTRODUCTION

COVID-19, the illness caused by the severe acute respiratory syndrome coronavirus 2 (SARS-CoV-2), has been diagnosed in over 80 million people worldwide as of the end of 2020, and over 1.8 million have died [1]. Although 81% of symptomatic individuals have relatively mild disease, 14% will develop severe disease characterized by dyspnea, hypoxia, or >50 percent lung involvement by imaging, with the remaining 5% developing critical disease characterized by respiratory failure, shock, and/or multiorgan dysfunction [2]. Severe and critical disease from COVID-19 is associated with advancing age (especially over 65 years), male gender, chronic lung disease, obesity, cardiovascular disease including hypertension, diabetes, and other chronic medical conditions.

Severe COVID-19 disease and death is, in part, mediated by rapid elevations of inflammatory cytokines including TNF-alpha, IL-1β, and IL-6 [3], leading to a cytokine release syndrome or “cytokine storm.” Accordingly, an attractive preventative approach to COVID-19 infection is to minimize cytokine release. Very long-chain omega-3s (DHA and EPA) found in fish oils have a plethora of biological activities including directly and indirectly modulating inflammatory responses and cytokine release. In non-COVID-19 ettings, higher intakes and levels of these omega-3s are associated with lower levels of circulating inflammatory cytokines[4-6], and intervention with fish oils reduces levels[7, 8]. EPA and DHA are precursors to a suite of inflammation-resolving mediators (IRMs; resolvins, maresins and protectins [9]) that actively regulate the resolution of acute inflammation. IRMs down-regulate cytokine production and promote a return to homeostasis via monocyte/macrophage-mediated uptake of debris, apoptosis of neutrophils, and clearing of microbes. Accordingly, higher intakes of EPA and DHA (which result in higher RBC EPA+DHA levels, hereafter called the Omega-3 Index, O3I [10, 11]) have been proposed to lower the risk for adverse outcomes from COVID-19 [12-18], and case reports suggesting benefit have been published [19, 20].

Given the profound public health concerns related to the current COVID-19 pandemic, modifiable risk factors for developing severe and critical complications are urgently needed, especially ones that may be nutritionally-based. Despite the known mechanisms by which IRMs and omega-3 fatty acids support the active, endogenous resolution of inflammatory mechanisms, to our knowledge no study has explored the relationship between omega-3 tissue levels and COVID-19 outcomes. The primary objective of this pilot study was to define the relationship between O3I and death from COVID-19. We hypothesize that a higher O3I is associated with lower risk for death in these patients.

## 2. METHODS

### 2.1. Subjects

We studied 100 patients hospitalized at Cedars-Sinai Medical Center from March 1, 2020 onwards with confirmed COVID-19 infection who met the criteria for inclusion, which were the availability of basic demographic data, clinical outcomes and an EDTA blood sample (drawn within 10 days of diagnosis) stored at −80°C at the Cedars-Sinai biorepository. If more than one sample was available, the first sample drawn after the time of diagnosis was utilized. We utilized sample remnant protocol Pro00036514 to obtain blood samples for this study.

Briefly, that protocol involved submission of a protocol to the Enterprise Information Service (EIS) team and staff members of the Biobank. After review and approval, the EIS/Biobank team pulled the samples and the data, de-identifying each sample by assigning a study-specific number. Using this protocol, we obtained a limited data and blood sample set to conduct this pilot investigation. The study was approved by the IRB of Cedars-Sinai Medical Center (STUDY-00000779).

### 2.2. Clinical outcomes

The primary outcome was death from COVID-19 infection.

### 2.3. Laboratory

Blood samples were thawed at the biorepository and one drop was placed on a dried blood spot collection card pre-treated with antioxidants to protect the fatty acids from degradation. The cards were then shipped overnight in batch to OmegaQuant Analytics (Sioux Falls, SD) for analysis of fatty acids and calculation of the Omega-3 Index[21]. Briefly, blood spots were transferred to a reaction vial and FA methyl esters (FAMEs) were generated using boron trifluoride in methanol by heating for 45 min at 100°C. FAMEs were extracted into hexane (after the addition of water) and analyzed using a GC2010 Gas Chromatograph (Shimadzu Corporation, Columbia, MD) equipped with a SP2560, 100-m column (Supelco, Bellefonte, PA). FAMEs were identified by comparison with a standard mixture (GLC, Nucheck Prep, Elysian, MN). Analysis was conducted using an internal-standard-based, three-point calibration curve to quantify levels of 24 FAMEs which were each expressed as a percent of total FAs. The O3I was calculated from the dried blood spot EPA+DHA value as described previously [21]. The analytical coefficient of variation for the O3I is <5%.

### 2.4. Power analysis

A sample size of 100 provided 80% power to detect a 10-fold difference in death risk between any quartile of the O3I and any other quartile, after adjusting for other variables and using an exploratory significance level of 0.10. We used a logistic regression model to test for a difference in death risk between any of the four quartiles of the O3I (4 group comparison), assuming that at least one quartile would be different than the others. We also had 80% power to detect a 3-fold difference in death risk between the highest quartile (Q4) of the O3I and all other (pooled) quartiles (Q1-3) after adjusting for other variables using a logistic regression model.

### 2.5. Statistical methods

The study sample is described using standard descriptive statistics (means and standard deviations, medians and ranges, and counts and percentages). Additional descriptive summary of the sample is provided by stratifying using quartiles of the O3I or by comparing Q4 to Q1-3. The distribution of age, sex and do not resuscitate (DNR) status by O3I quartiles or top 25%/bottom 75% was tested using an F-test (age) or Chi-squared test (sex, DNR status). The primary analysis examined the hypothesis that O3I is associated with risk for death. We first predicted death using unadjusted logistic regression models by quartiles of the O3I using a Chi-squared test to evaluate overall (4-group) association. We also predicted death by age, sex and DNR status to evaluate and confirm known associations between risk for death and these factors. Subsequently, significant association between O3I quartiles and death was tested using logistic regression and by comparing Q4 with Q1-3 in both unadjusted models and models adjusted for age and sex. To account for the small sample sizes present in this study, primary model results use Firth’s adjustment for small sample sizes in logistic regression using the logistf package in R [22]. We also report conventional logistic regression F-test results for comparison. Given the pilot/exploratory nature of this study, the sample size and our power analysis results, we set the statistical significance level to 0.10 with 2-tailed tests for all analyses.

## 3. RESULTS

### 3.1. Sample description

The study sample is described in **Table 1**. They were predominantly men, the mean age was over 70, and they were admitted between April and July 2020. Nearly 40% were under a DNR order, and 14% died during hospitalization. The average O3I was 5.09%, and the median was 4.75%.

**Table 1.** Descriptive statistics.

| Characteristics of sample | % (X/N) or Mean (SD; min,max) |
| --- | --- |
| Sex – Male | 59% (59/100) |
| Age | 72.5 (16.5; 25,100) |
| Month of data collection | April – 47% (47/100)<br>May – 25% (25/100)<br>June – 15% (15/100)<br>July – 13% (13/100) |
| Red blood cell EPA+DHA (% of RBC fatty acids; the Omega-3 Index, O3I) | 5.09% (1.62%; 2.87%, 13.79%) |
| Died | 14% (14/100) |
| DNR orders | 38% (38/100) |

Table 2 illustrates the association of the O3I by age, sex and DNR status. Higher O3I values were significantly related to older age, with the highest mean age in the third quartile (79.8 years). Men and those who were not DNR tended to have higher O3I values, but these trends were not statistically significant.

**Table 2.** Demographic profile of participants by category of the O3I.

| **Categorical by O3I Quartile** | **Age Mean (SD)** | **Sex - Male % (x/n)** | **DNR % (x/n)** |
| --- | --- | --- | --- |
| Q1 (O3I<4.0%) | 63.0 (13.3) | 64% (16/25) | 44% (11/25) |
| Q2 (4.0%<O3I<4.7%) | 71.04 (16.1) | 56% (14/25) | 28% (7/25) |
| Q3 (4.7<O3I<5.7%) | 79.8 (13.3) | 52% (13/25) | 52% (13/25) |
| Q4 (O3I≥5.7%) | 76.32 (12.72) | 64% (16/25) | 28% (7/25) |
| P-value<sup>1</sup> | **0.0015** | 0.77 | 0.20 |
| **Comparing O3I Q4 vs Q1-3** |  |  |  |
| Q1-3: O3I<5.7% | 71.25 (17.47) | 57.3% (43/75) | 41.3% (31/75) |
| Q4: O3I≥5.7% | 76.32 (12.72) | 64% (16/25) | 28% (7/25) |
| P-value<sup>1</sup> | 0.19 | 0.55 | 0.28 |

<sup>1</sup>F-test (age and O3I) or Chi-square test (sex/DNR and O3I).

### 3.2. Unadjusted analyses

Relationships between omega-3 status and fatal outcomes are shown in **Table 3**. As expected, older patients and those under a DNR order were more likely to die. In the unadjusted model (and focusing on the small-sample size p-values), there was a significant difference in risk for death across quartiles of the O3I (p=0.047), with those in the highest O3I quartile (Q4) having an odds ratio (OR) of 0.39 (p=0.34) relative to Q1. In an unadjusted comparison between Q4 and Q1-3, the OR for death in Q4 was 0.28 (p=0.11). For comparison, the OR for death from COVID-19 per 1-decade greater age (OR=1.33, p=0.14) was of about the same magnitude and significance as for being in O3I Q1-3 vs Q4. The unadjusted relative risk for death for those in the lower three quartiles was 4.3 (i.e., 17.3%/4.0%). In Q4, there was one death (a 66-year-old male under a DNR order), whereas there were 13 deaths among the 75 patients in Q1-3. The higher OR in Q3 was largely accounted for by age as the mean age in Q3 was the highest of all. The OR in Q3 was markedly attenuated in the adjusted analysis (**Table 4**).

**Table 3.** Unadjusted associations of the Omega-3 Index and demographic variables with death.

| Risk factor | Death % (x/N) | Unadjusted Models |  |  |
| --- | --- | --- | --- | --- |
|  |  | OR (95% CI)<sup>1</sup> | Firth's test P-value<sup>1</sup> | F-test P-value<sup>2</sup> |
| **Sex** |  |  |  |  |
| Female | 14.6% (6/41) | 1.00 |  |  |
| Male | 13.6% (8/59) | 0.90 (0.30, 2.83) | 0.85 | 0.88 |
| **Age** |  |  |  |  |
| Per decade | -- | 1.33 (0.92, 2.08) | 0.14 | 0.13 |
| **DNR** |  |  |  |  |
| Yes | 34.2% (13/38) | 21.76 (4.90, 206.44) | **<0.0001** | **0.001** |
| No | 1.6% (1/62) |  |  |  |
| **Categorical by O3I Quartile** |  | Overall model F-test | **0.047** | **0.03** |
| Q1 (O3I<4.0%) | 12.0% (3/25) | 1.00 |  |  |
| Q2 (4.0%<O3I<4.7%) | 8.0% (2/25) | 0.68 (0.11, 3.86) | 0.66 | 0.64 |
| Q3 (4.7<O3I<5.7%) | 32.0% (8/25) | 3.13 (0.82, 14.30) | 0.10 | 0.10 |
| Q4 (O3I≥5.7%) | 4.0% (1/25) | 0.39 (0.04, 2.61) | 0.34 | 0.32 |
| **Comparing O3I Q4 vs Q1-3** |  |  |  |  |
| Q1-3: O3I<5.7% | 17.3% (13/75) | 1.00 |  |  |
| Q4: O3I≥5.7% | 4.0% (1/25) | 0.28 (0.03, 1.26) | 0.11 | 0.13 |

Bold and italics is used for p-values less than the significance level of 0.10. <sup>1</sup> Using the Firth's modified score procedure for small sample sizes. <sup>2</sup> Standard F-test from Logistic Regression ignoring small sample sizes.

**Table 4.**
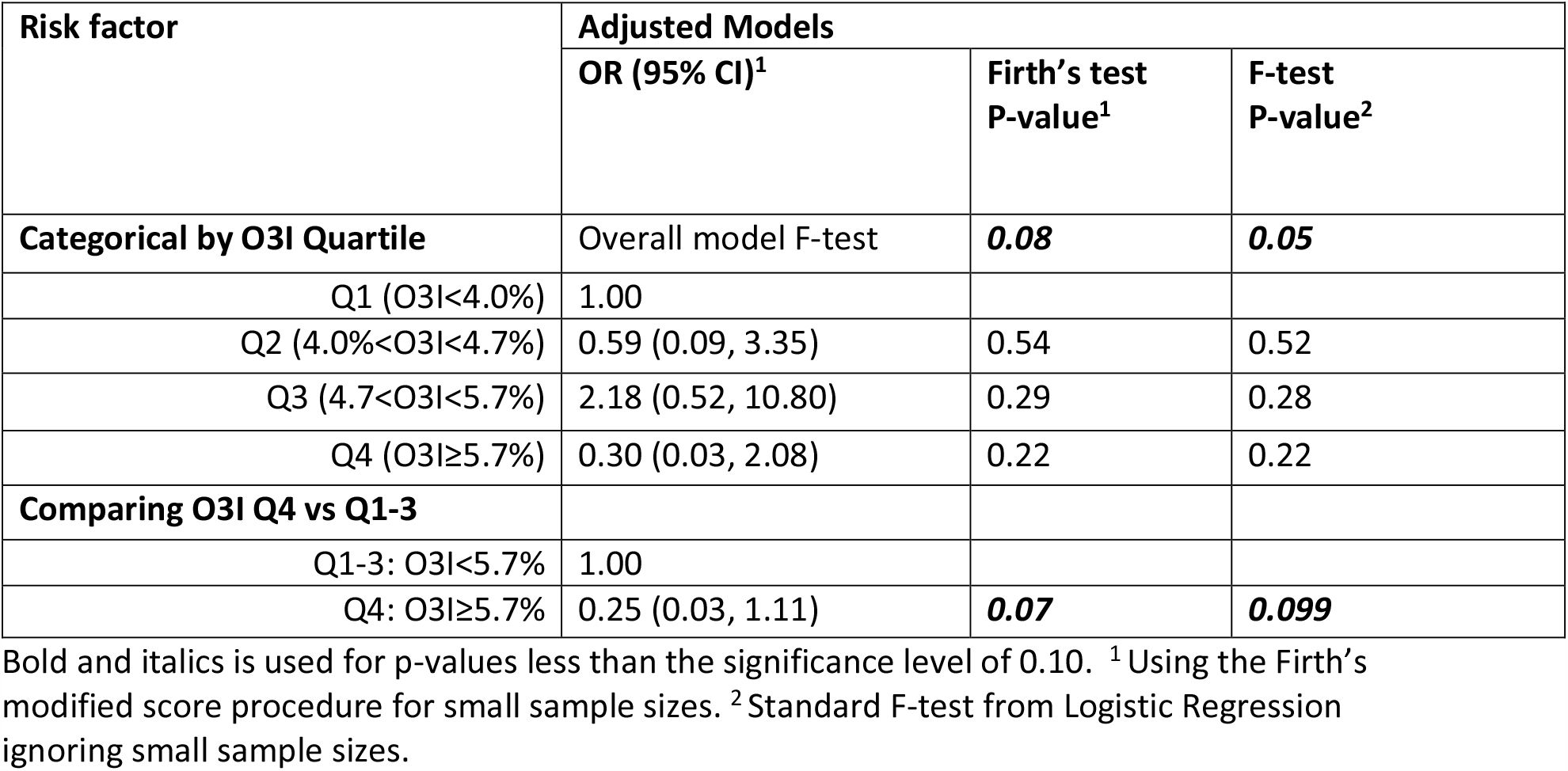
Associations of the Omega-3 Index with death adjusted for age and sex.

### 3.3. Adjusted analyses

In models adjusted for age and sex (**Table 4**), the overall relationship across quartiles became somewhat weaker (i.e., from p=0.047 to 0.08), and although the OR at Q4 (vs Q1) remained the same (0.3), the p-value decreased somewhat (from 0.32 to 0.22). Comparing Q4 to Q1-3 in the adjusted analysis, the OR increased (from 0.20 to 0.25), but the p-value decreased from 0.13 to 0.07. In other words, those in Q4, with an O3I >5.7%, were 75% less likely to die from COVID-19 compared with those with an O3I<5.7%.

## 4. DISCUSSION

The 2020 COVID-19 pandemic has had devasting effects on morbidity and mortality worldwide. While vaccines may soon slow the spread and drugs may help diminish the adverse effects of infection, preventative measures – ideally ones that are cheap, safe, and readily accessed by large numbers of people – that modulate the most severe disease outcomes are desperately needed. Some evidence is accumulating for a potential benefit of vitamin D [23], and here we examined another nutrient, omega-3 fatty acids, which like vitamin D, have multiple anti-inflammatory effects and may also reduce risk for adverse COVID-19 outcomes.

In this pilot study we compared the associations between the O3I and risk for death from COVID-19 in 100 patients. At our exploratory significance threshold of 0.10, we found (in age and sex-adjusted models) that those patients with an O3I at 5.7% or greater were at about 75% lower risk for death compared with those below that value (p=0.07). These findings suggest that a relationship may indeed exist, but larger studies are clearly needed.

This study used the O3I as a biomarker of omega-3 FA status. This RBC-based metric has advantages, particularly in the acute hospitalization setting. This is because, much like a hemoglobin A1C versus plasma glucose, the O3I is a better long-term reflection of tissue omega-3 levels versus plasma omega-3 levels [23, 24], and it would thus less affected by an acute change in omega-3 intake as might happen with hospitalization for an acute illness [25]. The O3I has been validated [21] and used in several interventional [10] and prospective cohort studies such as the Framingham Heart Study [26] and the Women’s Health Initiative Memory Study [27]. The O3I is also easily modified by increasing the intake of oily fish (e.g., salmon, herring, mackerel, albacore tuna, etc.) which are rich in EPA and DHA. Dietary supplements of omega-3 will also raise levels [11]. The average O3I in this study was 5.1% which is similar to that seen other US-based studies [28, 29].

Multiple randomized clinical trials (RCTs) are currently (as of January 2021) underway testing the hypothesis that treatment with omega-3 fatty acids (EPA and DHA) will have beneficial effects on a variety of aspects of COVID-19 infection. Although their outcomes are not yet known, there are compelling scientific reasons to expect that these studies will be positive (and these same reasons formed the foundation for the present study). The data supporting a possible beneficial role for omega-3 fatty acids in COVID-19 infection come from past epidemiological, interventional, therapeutic, and basic science studies. For example, in the Framingham Offspring study [30], the O3I was inversely associated with 10 separate inflammatory biomarkers (e.g., CRP, IL-6, ICAM-1, LpPLA2, TNF receptor 2, and osteoprotegerin). In intervention studies [7], EPA supplementation alone (3g/d for 10 weeks) significantly reduced the expression of TNFa from LPS-stimulated monocytes as did a similar dose of DHA which, in addition, lowered IL-6 and MCP-1. EPA+DHA supplements had similar effects [31]. Meta-analyses of multiple RCTs confirmed that treatment with omega-3 fatty acids routinely lowers cytokine levels [32-34]. More important than studies of effects on intermediate markers are clinical findings from RCTs. Langlois et al. (2018) summarized the results of 12 RCTs of omega-3 treatment in 1280 intensive-care-unit patients with acute respiratory distress syndrome. There was a significant improvement in measures of blood oxygenation in the treated patients and strong trends (p≤0.08) for reduced ICU length of stay and duration of mechanical ventilation. Overall mortality, hospital length of stay and infectious complications were unaffected.

As noted earlier, the potential mechanisms underlying these actions are multiple (**Figure**). EPA/DHA are substrates for the production of IRMs which cannot be made if the parent compounds are not present. Examples of some of the functions of IRMs were summarized by Calder as, “Resolvin E1, resolvin D1 and protectin D1 all inhibit trans-endothelial migration of neutrophils, so preventing the infiltration of neutrophils into sites of inflammation; resolvin D1 inhibits IL-1β production and protectin D1 inhibits TNF-α and IL-1β production.”[35] Higher EPA/DHA levels reduce arachidonic acid (the omega-6 cousin of EPA) membrane levels [36] for the production of some pro-inflammatory oxylipins (certain prostaglandins and leukotrienes). Quite independently of the synthesis of these mediators, the presence of EPA/DHA in inflammatory cells blocks the activation of the key pro-inflammatory transcription factor, nuclear factor kappa B thus retarding the entire intracellular inflammatory cascade [37, 38].

**Figure.**
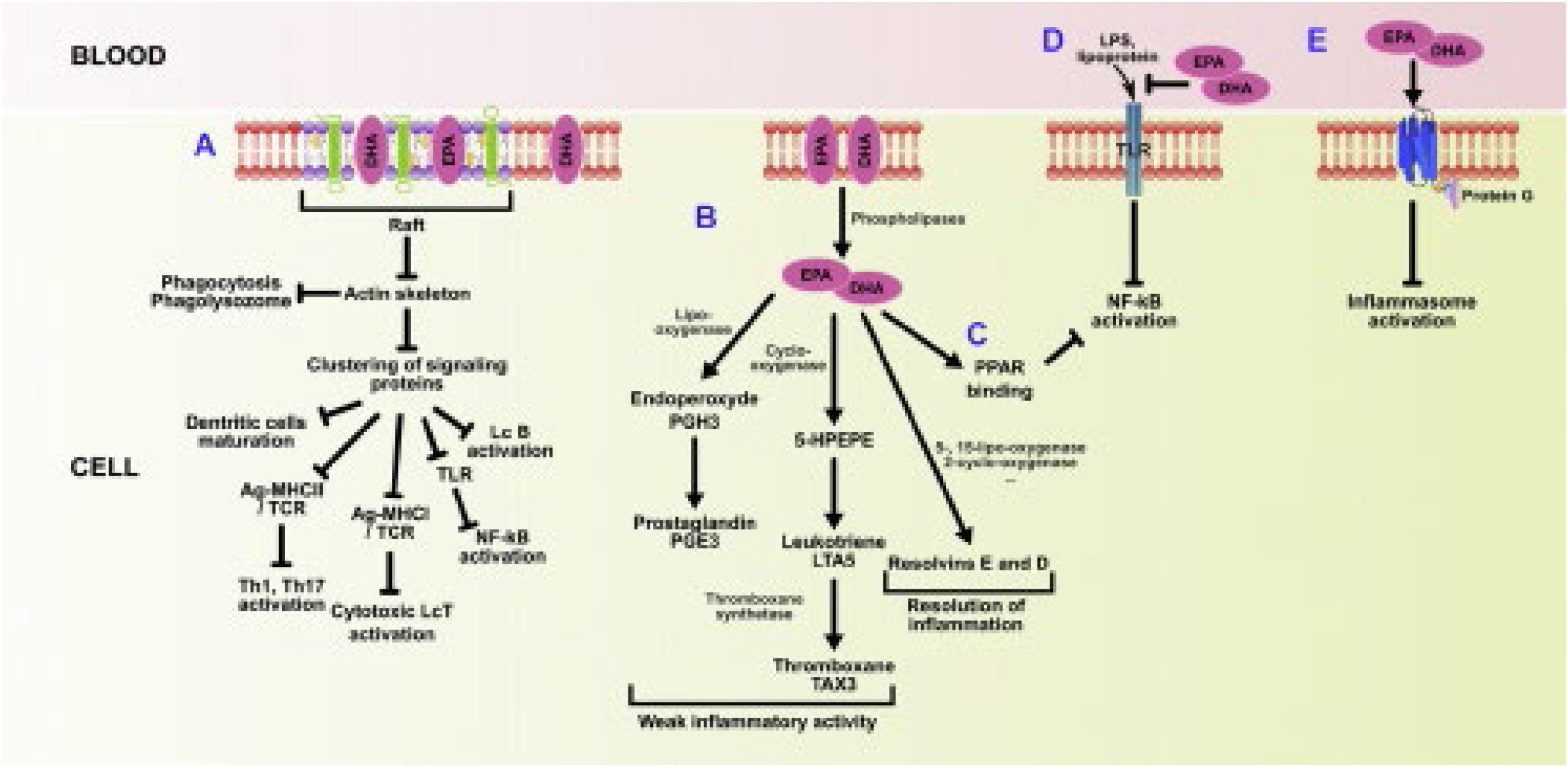
Potential mechanisms underlying the anti-inflammatory actions of EPA and DHA. A) In cell membranes, disruption of rafts which impedes signaling and thereby affects Th1, Th7, and cytotoxic T cells, B activation, and TLR clustering; B) in cells, synthesis of arachidonic acid-derived leukotrienes, prostaglandins, and thromboxanes is reduced and replaced by EPA-derived isomers with weaker inflammatory effects; and resolvins are produced to accelerate the resolution of inflammation; C) in cells, binding of EPA and DHA to PPAR inhibits NF-kB activation; D) in serum, reduced lipopolysaccharide binding to toll-like receptor 4; and E) in serum, inhibition of inflammasome activation. Reprinted with permission from Elsevier and Husson et al.[6]

This effect is downstream from the action of these fatty acids on membrane G-protein coupled receptors GPR40 and GPR120 and nuclear peroxisome proliferator-activated receptors (reviewed in [39]). Finally, EPA/DHA insert into cell membrane phospholipids and disrupts lipid rafts so as to disassemble surface receptors thereby blocking incoming inflammatory signals [40]. All of these actions together result in a muted “cytokine storm” (i.e., rapid over-expression of cytokines) which, in alveolar macrophages, can rapidly result in death from COVID-19.

### 4.1. Limitations

Given the pilot nature of this exploratory study, a number of limitations are acknowledged. Firstly, the sample size was small. Secondly, the limited resources and resulting access to the full electronic medical record for more detailed chart review significantly reduced the amount of potentially relevant information on comorbidities and other demographic data besides age and sex (e.g., BMI). Although data on maximal interventions applied during hospitalization were available, the reasons why any given patient was administered a given treatment are not known and could obviously have been influenced by external factors (e.g., ventilator availability) or internal factors (e.g., a DNR order). Accordingly, such information was deemed to be of little utility in this pilot study. The population of patients in this study had an O3I that was typical of the US [28], which unfortunately means that levels were generally low. Future studies should endeavor to include patients with a wider range of O3I to examine these relationships more clearly.

### 4.2. Conclusions

Given the profound public health concerns related to the current COVID-19 pandemic, modifiable risk factors for developing severe and critical complications are urgently needed. Despite the known mechanisms by which IRMs and omega-3 fatty acids support the active, endogenous resolution of inflammatory mechanisms, to our knowledge this is the first study that has explored the relationship between omega 3 tissue levels and the most severe COVID-19 outcome, death. Larger studies are urgently needed to confirm these findings. If an association is confirmed with a larger sample size, then this would lay the groundwork for testing the effects of increased oily fish intake and/or an inexpensive, safe, and widely available dietary supplement (DHA/EPA capsules) to optimize outcomes during this public health crisis.

## Data Availability

The data that support the findings of this study are available from the corresponding author upon reasonable request.

